# Diagnostic Accuracy of Nanopore Sequencing for Detecting *Mycobacterium tuberculosis* and Drug-Resistant Strains: A Systematic Review and Meta-Analysis

**DOI:** 10.1101/2024.06.05.24308511

**Authors:** Timothy Hudson David Culasino Carandang, Dianne Jaula Cunanan, Gail S. Co, John David Pilapil, Juan Ignacio Garcia, Blanca I. Restrepo, Marcel Yotebieng, Jordi B. Torrelles, Kin Israel Notarte

## Abstract

Tuberculosis (TB), caused by *Mycobacterium tuberculosis* (MTB) infection, remains a significant public health threat. The timeliness, portability, and capacity of nanopore sequencing for diagnostics can aid in early detection and drug susceptibility testing (DST), which is crucial for effective TB control. This study synthesized current evidence on the diagnostic accuracy of the nanopore sequencing technology in detecting MTB and its DST profile. A comprehensive literature search in PubMed, Scopus, MEDLINE, Cochrane, EMBASE, Web of Science, AIM, IMEMR, IMSEAR, LILACS, WPRO, HERDIN Plus, MedRxiv, and BioRxiv was performed. Quality was assessed using the Quality Assessment of Diagnostic Accuracy Studies-2 tool. Pooled sensitivity, specificity, predictive values (PV), diagnostic odds ratio (DOR), and area under the curve (AUC) were calculated. Thirty-two studies were included; 13 addressed MTB detection only, 15 focused on DST only, and 4 examined both MTB detection and DST. No study used Flongle or PromethION. Seven studies were eligible for meta-analysis on MTB detection and five for DST; studies for MTB detection used GridION only while those for DST profile used MinION only. Our results indicate that GridION device has high sensitivity [88.61%; 95% CI (83.81–92.12%)] and specificity [93.18%; 95% CI (85.32–96.98%)], high positive predictive value [94.71%; 95% CI (89.99–97.27%)], moderately high negative predictive value [84.33%; 95% CI (72.02–91.84%)], and excellent DOR [107.23; 95% CI (35.15–327.15)] and AUC (0.932) in detecting MTB. Based on DOR and AUC, the MinION excelled in detecting pyrazinamide and rifampicin resistance; however, it underperformed in detecting isoniazid and ethambutol resistance. Additional studies will be needed to provide more precise estimates for MinION’s sensitivity in detecting drug-resistance, as well as DOR in detecting resistance to pyrazinamide, streptomycin, and ofloxacin. Studies on detecting resistance to bedaquiline, pretomanid, and linezolid are lacking.

## INTRODUCTION

Tuberculosis (TB), caused by *Mycobacterium tuberculosis* (MTB) infection, remains a significant public health threat, with an estimated 10 million new infections annually^1^. In 2022, it was the second leading cause of mortality among infectious diseases after COVID-19^2^. Though affecting all countries and age groups, TB is most prevalent in low- and middle-income countries (LMICs), with almost half of the cases occurring in mainly eight countries (Bangladesh, China, India, Indonesia, Nigeria, Pakistan, Philippines, and South Africa)^2^. Early detection and diagnosis can dramatically affect treatment outcomes and prognosis, preventing the development of drug resistance. Yet, some diagnostic challenges present, such as lack of point-of-care (POC) tests that can be accessed in remote areas and the long turnaround time of the culture, which to this date remains the gold standard for diagnosis^3^.

Nucleic acid amplification tests (NAAT) such as GeneXpert^®^ MTB/RIF (Xpert) have improved the detection and management of TB. Due to its timeliness, high sensitivity, and high specificity compared to acid-fast bacilli (AFB) smears and cultures, the Xpert has gained widespread use, particularly in diagnosing multi-drug resistant (MDR) TB, as well as TB in people living with HIV (PLWH). Numerous studies have been performed evaluating Xpert’s utility^4,5^, prompting policy recommendations from the World Health Organization (WHO)^6^. While Xpert offers rapid diagnosis, its high laboratory cost, encompassing consumables, annual calibrations, and maintenance, restricts its use in resource-limited settings where access to NAAT technologies might be inconsistent.^4,7,8^

Recent developments in third-generation sequencing technology, such as Oxford Nanopore Technologies sequencing platforms, have shown several advantages over other diagnostic tools used for TB. In this context, nanopore sequencing is being applied in full-length transcript detection and base modification detection focused on rapid clinical diagnoses. MTB is an ideal candidate for analysis since nanopore sequencing excels in the investigation of complex genomic loci and large repetitive elements^9^. The ability of nanopore sequencing to generate long read lengths and conduct real-time analysis offers rapid detection and comprehensive profiling of drug resistance, leading to prompt and suitable treatment strategies, minimizing treatment-related costs, and improving patient outcomes. Its portability and capacity to directly sequence nucleic acid molecules simplify the workflow and allow on-site sequencing, improving access to quality diagnostics even in LMICs^10^. This technology depends on nanoscale protein pores working as biosensors of changes in ionic current as the DNA or RNA is translocated into each pore^11^. Each DNA or RNA base has a fixed size that blocks current flow as the strand is translocated, registering a unique and interpretable alteration in current.

To date, there is no systematic review nor meta-analysis of existing studies on the diagnostic accuracy of nanopore sequencing for TB detection and DST. To address this important knowledge gap, this study aims to appraise and synthesize current evidence on the sensitivity, specificity, negative predictive value (NPV), positive predictive value (PPV), diagnostic odds ratio (DOR) and area under the curve (AUC) of nanopore sequencing technology in detecting MTB and its drug-resistant patterns.

## METHODS

*Study protocol and registration* - The study protocol was submitted to The International Prospective Register of Systematic Reviews (PROSPERO) (ID: CRD42024518764) and reported in compliance with the PRISMA 2020 guidelines^12^.

*Search strategy* - Literature searches were independently conducted by two different co-authors using the following databases: PubMed, Scopus, MEDLINE, Cochrane, EMBASE, Web of Science, AIM, IMEMR, IMSEAR, LILACS, WPRO, HERDIN Plus, MedRxiv and BioRxiv. Separate comprehensive searches were done for studies about MTB detection and drug susceptibility. As the first nanopore sequencing platform, namely minION, was first commercialized in 2014, our search range was narrowed down to English articles published from January 2014 until February 2024^13^. The selected reference list of each article was also screened to identify studies relevant to this review. The following search terms were used: “*Mycobacterium tuberculosis*” OR “*M. tuberculosis”* AND “nanopore sequencing” OR “nanopore-based sequencing” OR “Oxford nanopore” OR “nanopore technology” OR “Flongle” OR “minION” OR “gridION” OR “promethION”. The combination of these search terms is detailed in the supplementary materials.

*Study selection* - All studies that compare nanopore sequencing technology to any conventional diagnostic test or reference standard in MTB detection and/or DST on clinical specimens was included except reviews, meta-analyses, commentary papers, case reports, conference summaries, animal experiments, and non-English literature. Two researchers independently screened the titles and abstracts. Disagreements were resolved by a third researcher in a blinded manner.

*Data extraction* - Basic diagnostic accuracy parameters such as true positive, false positive, true negative, and false negative were extracted using a 2 X 2 table. Other relevant data included the following: year, country, research type, sample size, type of nanopore sequencing technology, comparator group, reference standard, sample type, and average age of patients. Research type was determined using the classification algorithm by Mathes & Pieper^14^. Data extraction was carried out independently by two researchers.

*Risk of bias assessment* - The risk of bias in diagnostic accuracy was assessed using the QUADAS-2 tool^15^. One researcher independently evaluated all included studies. Risk of bias assessment was carried out independently by two researchers.

*Statistical analysis* - Pooled sensitivity, specificity, NPV, PPV, and DOR were reported with 95% confident interval (CI). Heterogeneity was quantified using the inconsistency value (I2); where I2 >50% is considered as a remarkable heterogeneity, which warranted subgroup analysis or meta-regression to identify sources of heterogeneity. All pooled effect sizes were determined using a random effects model to account for both within- and between-studies variance. Receiver operating characteristic (ROC) curves were plotted to assess the overall accuracy of nanopore sequencing; there was a high diagnostic value if the AUC values exceeded 0.9. Subgroup analyses and meta-regression were not performed since there were < 10 studies included in the meta-analysis. Deek’s test for publication bias was performed if there were ≥ 10 eligible studies with computable diagnostic odds ratios^16^. Statistical significance was set at *p* <0.05. Analyses were performed using the “*mada*” and “*meta*” packages of R statistical software vr. 4.3.3 and IBM SPSS vr. 26^17^.

## RESULTS

### Characteristics of included studies

Comprehensive search across multiple databases yielded a total of 1,073 records about MTB detection and 226 records about MTB drug susceptibility. The full search strategies were summarized in **Supplementary Tables 1and 2**. Following a thorough screening process, 17 studies about MTB detection were included while 19 studies about MTB drug susceptibility were selected (**Fig. 1**).

**Figure 1.**
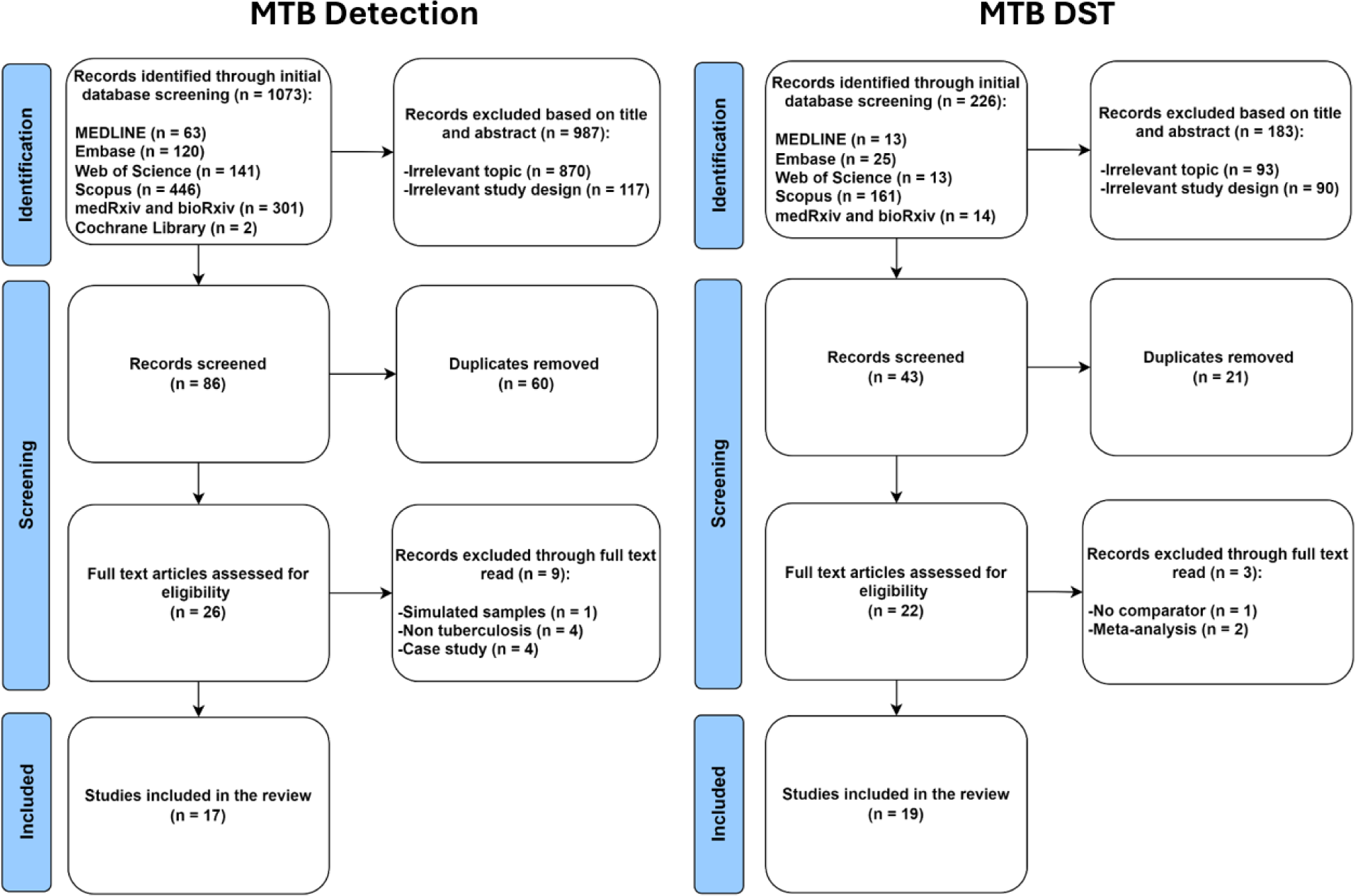
PRISMA flow diagram of this study selection process.

To facilitate our analysis, studies were classified into two categories: those focusing on the detection of MTB (**Table 1**) and those focusing on the detection of DR-MTB (**Table 2**). Among the 32 articles, 13 articles related to MTB detection only^18-30^,15 articles focused on DR-MTB detection only^31-45^, while 4 articles examined both^46-49^.

**Table 1.**
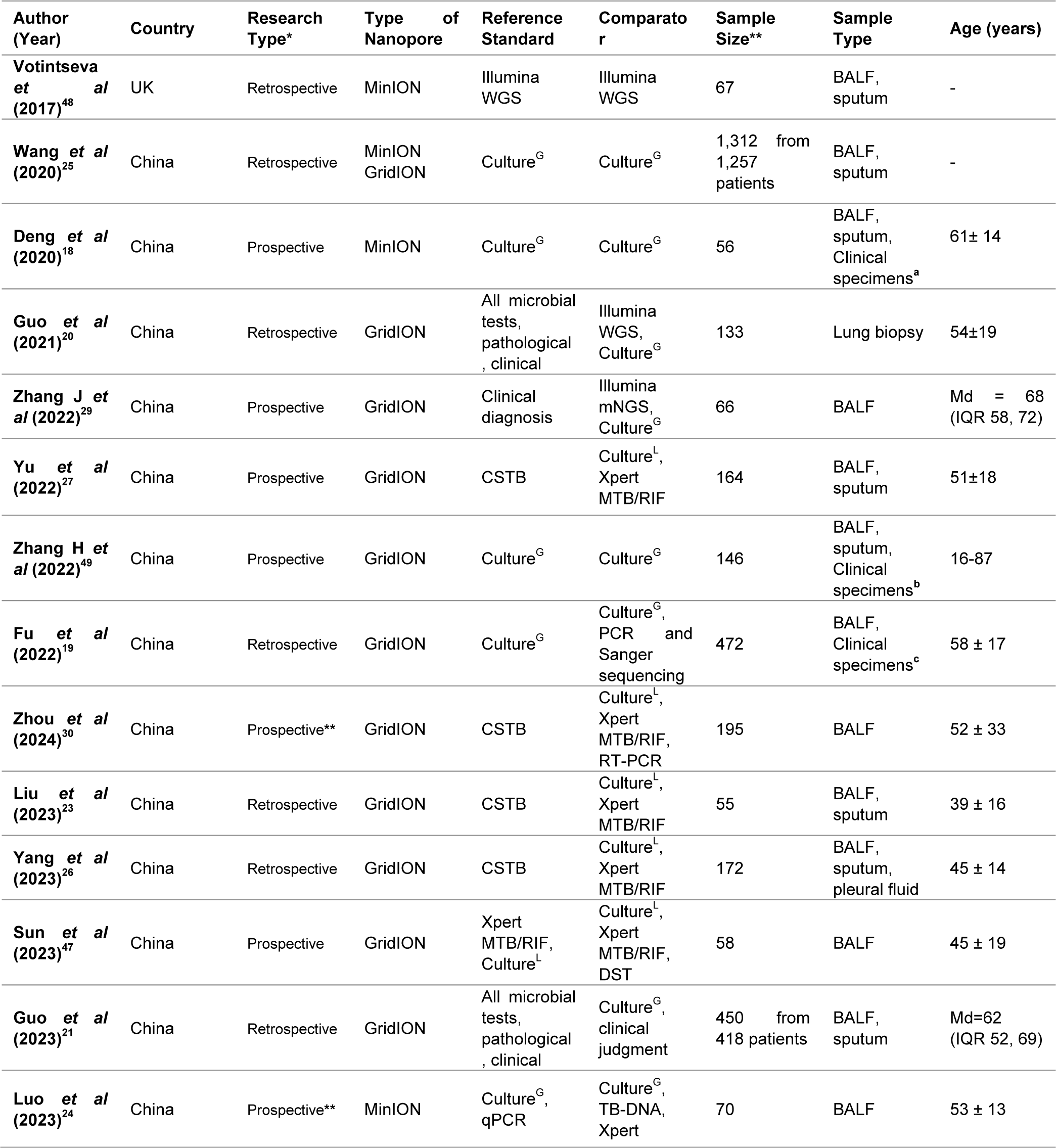
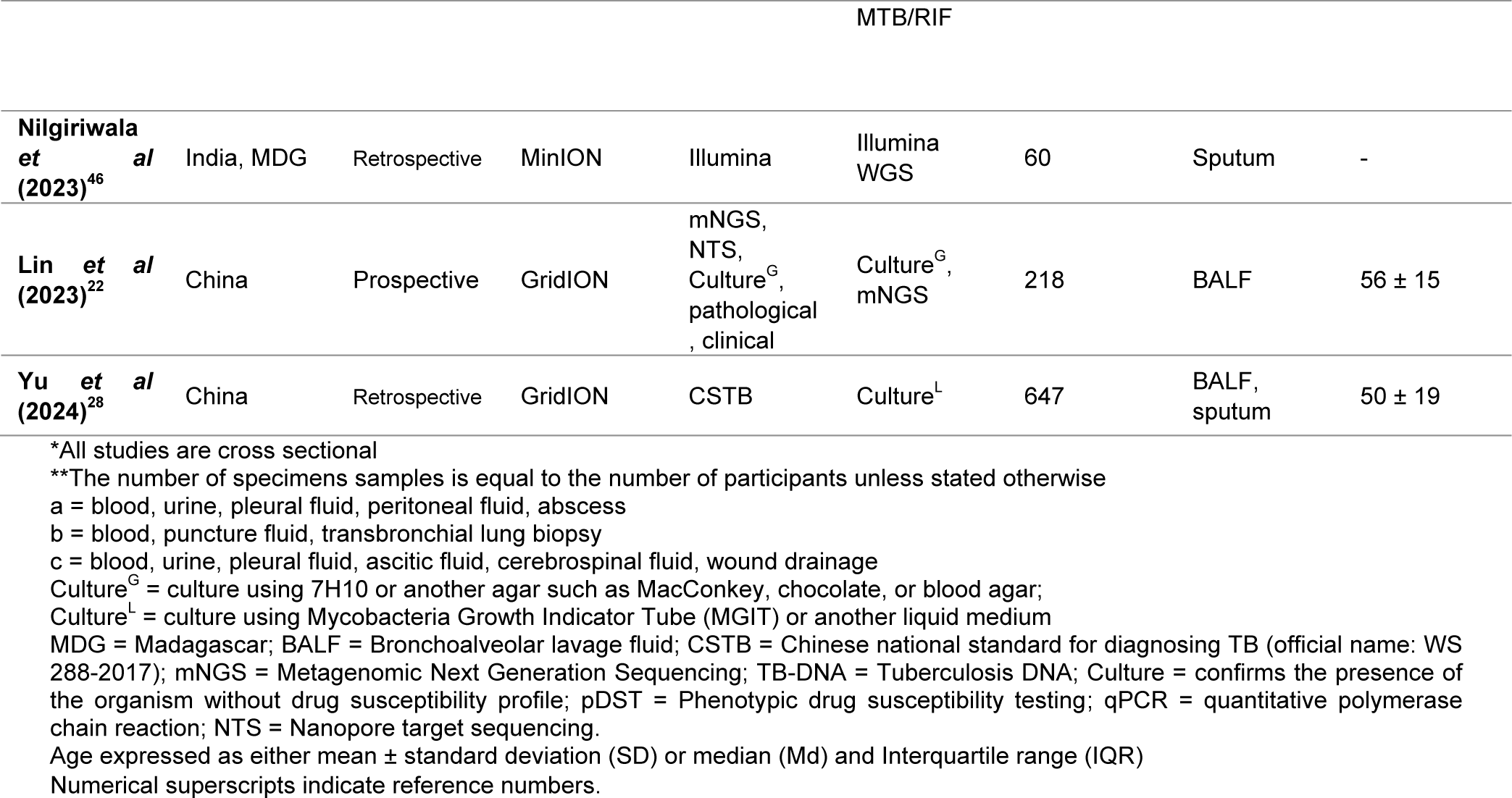
Characteristics of included studies on detection of MTB.

**Table 2.** Characteristics of included studies on detection of DR-MTB.

| Author (Year) | Country | Research Type* | Type of Nanopore | Reference Standard | Comparator | Sample Size | Sample Type | Age (years) |
| --- | --- | --- | --- | --- | --- | --- | --- | --- |
| Votintseva et al (2017)<sup>48</sup> | UK | Retrospective | MinION | pDST | Illumina WGS | 67 | BALF, sputum | - |
| Smith et al (2020)<sup>39</sup> | USA | Prospective | MinION | pDST | Illumina, pDST | 431 | DNA extracts^ | - |
| Cervantes et al (2020)<sup>32</sup> | USA | Retrospective | MinION | Illumina WGS | Illumina WGS | 6 | Sputum, DNA extracts^ | - |
| Chan et al (2020)<sup>33</sup> | South Africa | Prospective | MinION | Illumina WGS | Illumina WGS | 23 | Sputum, bronchial aspirate, biopsy<sup>a</sup> | - |
| Tafess et al (2020)<sup>40</sup> | Hongkong Ethiopia | Prospective | MinION | pDST | Illumina WGS | 163 | DNA extracts^ | - |
| Cabibbe et al (2020)<sup>31</sup> | Italy | Retrospective | MinION | Illumina WGS | Illumina WGS | 104 | DNA extracts^ | - |
| Gliddon et al (2021)<sup>34</sup> | United Kingdom | Retrospective | MinION | pDST | Illumina WGS, pDST | 34 | Sputum | Md=36 (IQR, 31-42) |
| Mariner-Llicer et al (2021)<sup>37</sup> | Spain | Prospective | MinION | Illumina WGS | Illumina WGS | 15 | Sputum, culture | - |
| Yu et al (2021)<sup>44</sup> | Taiwan | Retrospective | MinION | pDST | - | 50 | DNA extracts^ | - |
| Zhang H et al (2022)<sup>49</sup> | China | Prospective | GridION | Culture<sup>G</sup> | Culture<sup>G</sup> | 146 | BALF, sputum, Clinical specimens<sup>b</sup> | 16-87 |
| Gómez-González et al (2022)<sup>35</sup> | UK | Retrospective | MinION | pDST | Illumina WGS | 10 | DNA extracts^ | - |
| Yu et al (2022)<sup>45</sup> | Taiwan | Retrospective | MinION | pDST | - | 50 | DNA extracts^ | - |
| Zhao et al (2022)<sup>41</sup> | China | Prospective | MinION | Culture<sup>G</sup> | PCR with Sanger sequencing | 20 | Sputum | 40 ± 11 |
| Sun et al (2023)<sup>47</sup> | China | Prospective | GridION | pDST | Culture, Xpert MTB/RIF, pDST | 58 | BALF | 45 ± 19 |
| Hall et al (2023)<sup>36</sup> | MDG, South Africa, England | Prospective | MinION, GridION | pDST | Culture<sup>G</sup>, Illumina WGS | 208 | DNA extracts^ | - |
| Nilgiriwala et al (2023)<sup>46</sup> | India, MDG | Prospective | MinION | Illumina WGS | Illumina WGS | 60 | Sputum | - |
| **Murphy et al (2023)<sup>38</sup>** | USA | Prospective | MinION, GridION | pDST | RT-PCR, Illumina WGS | 127 | BALF, sputum, biopsy<sup>a</sup> | - |
| **Zhao et al (2024)<sup>42</sup>** | Taiwan | Prospective | GridION | pDST | Culture<sup>G</sup> | 297 | BALF, sputum, clinical specimens<sup>c</sup> | 55 ± 18 |
| **Liu et al (2024)<sup>43</sup>** | China | Prospective | GridION | pDST + Xpert MTB/RIF | - | 184 | BALF, sputum, hydrothorax, ascites, pus | Md=48 (IQR, 37-57) |

\*All studies are cross-sectional
^DNA extracts refer to DNA extractions from MTB-positive clinical isolates with unspecified source
a = lymph node biopsy, lung biopsy
b = blood, puncture fluid, transbronchial lung biopsy
c = plasma, cerebrospinal fluids, abscess, serous cavity effusions, and urine
Culture<sup>G</sup> = culture using 7H10 or another agar such as MacConkey, chocolate, or blood agar;
Culture<sup>L</sup> = culture using Mycobacteria Growth Indicator Tube (MGIT) or another liquid medium
MDG = Madagascar; BALF = Bronchoalveolar lavage fluid; mNGS = Metagenomic Next Generation Sequencing; Culture = confirms the presence of the organism without drug susceptibility profile; pDST = Phenotypic drug susceptibility testing; RT-PCR = reverse-transcriptase polymerase chain reaction; WGS = Whole Genome Sequencing
Age expressed as either mean ± standard deviation (SD) or median (Md) and Interquartile range (IQR)
Numerical superscripts indicate reference numbers.

### Detection of *MTB*

Out of the 17 articles, nine were retrospective cross-sectional while eight were prospective cross-sectional. GridION was predominantly used in these studies (12 out of 17), followed by minION alone (4 out of 19), then the combination of both sequencing platforms (1 out of 19). No study used Flongle or PromethION. Included studies were published between 2017 and 2024 (**Table 1**).

### Risk of bias and applicability assessment for detection of MTB

Separate assessments of the risk of bias and applicability concerns using the QUADAS-2 tool were performed for studies on MTB detection (**Figure 2**) and drug susceptibility (**Figure 4**). Mainly, patient selection is the top source of bias and applicability concerns. In the 17 MTB detection studies selected, only one explicitly described the sampling strategy, consisting of consecutive enrollment^47^ (**Table 1**). Further, nine of the selected studies were focused on general infections, lower respiratory tract infections, and pneumonia, and thus, their MTB results derived from secondary analyses; hence, the heightened applicability concerns. Additionally, among the 17 MTB detection studies, nine adopted a retrospective cross-sectional design (**Figure 2**), increasing the risk of bias concerning the evaluated index test, reference test, and the flow of timing in these studies.

**Figure 2.**
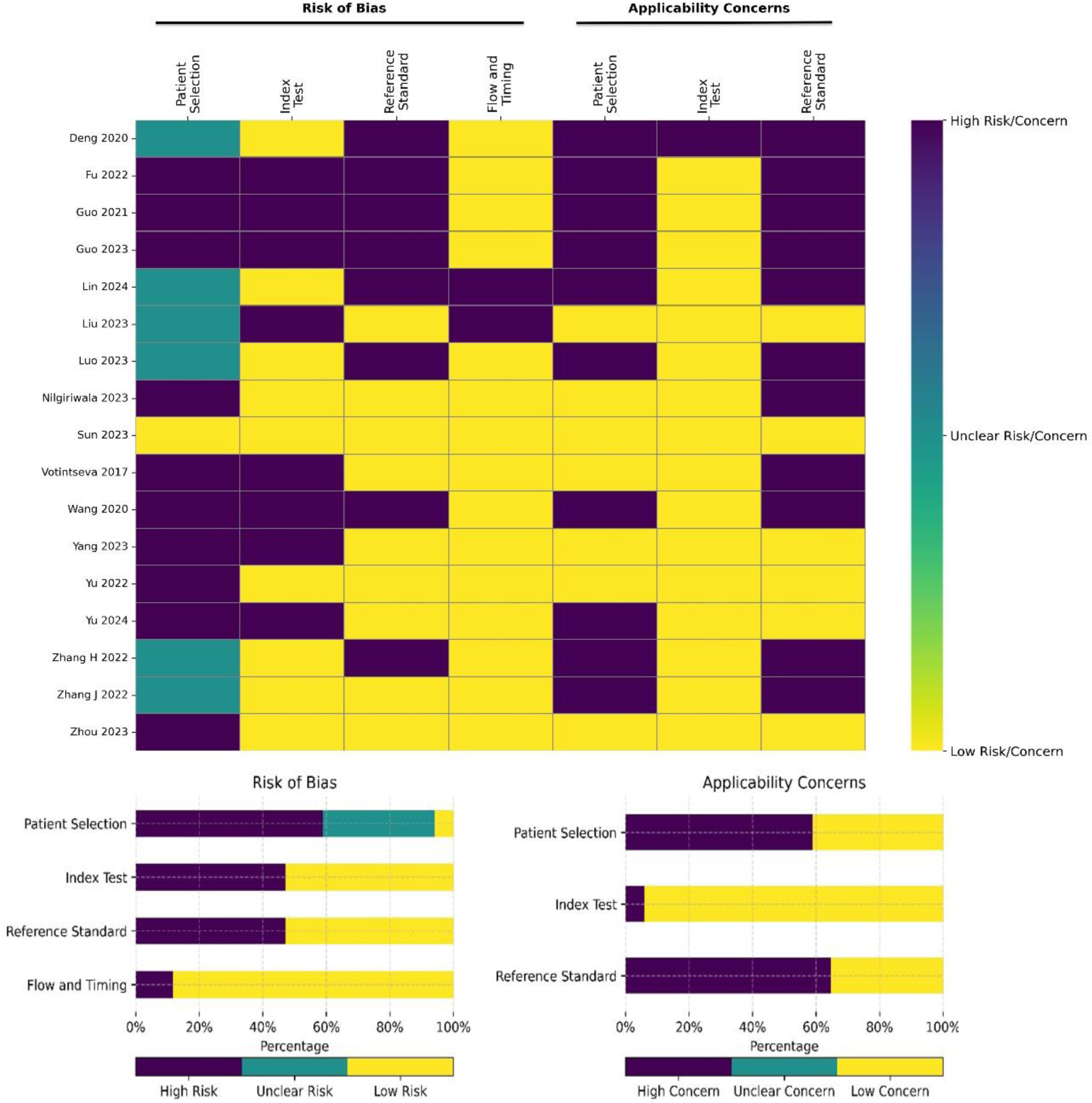
Heat map showing risk of bias and applicability concerns among the selected 17 studies focusing on the detection of MTB.

### Meta-analysis of MTB detection studies

*Heterogeneity test.* Our results showed that the logit-transformed sensitivity did not correlate with logit transformed false positive rate (1-specificity) (Spearman correlation -0.486 (*p* = 0.356)). Our analysis revealed no threshold effect among the included studies. Higgin’s I2 of the diagnostic measures ranged from 63% -88%.

*Evaluation of diagnostic accuracy.* The meta-analysis for detecting MTB included seven studies, with 1,709 specimens from 1,741 adult participants. Our results indicate that the diagnostic sensitivity of nanopore sequencing in detecting MTB ranged from 75% to 95%, while its specificity varied between 81% and 100%. Nanopore sequencing had a pooled sensitivity of 88.61% (95% CI 83.81–92.12%, I2 = 63%), and a pooled specificity of 93.18% (95% CI: 85.32– 96.98%, I2 = 77%) (**Figure 3**). Its PPV ranged from 81% to 100%, while its NPV varied between 67% and 99%. The pooled PPV value was 94.71% (95% CI 89.99–97.27%, I2 = 65%), while its pooled NPV was computed to be 84.33% (95% CI: 72.02–91.84%, I2 = 88%) (**Figure 3**). Our results also indicated that AUC was excellent [AUC = 0.932 (SE = 0.03)], with a remarkable wide DOR range, where values varied from 13 to 1,311. Pooled DOR was determined to be 107.23 (95% CI 35.15–327.15), with moderate heterogeneity (I2 = 65%) (**Figure 3**). The summary receiver operating characteristic (SROC) curve for nanopore sequencing in MTB detection is shown in **Figure 3**. Youden’s J index was calculated to be 0.82 (**Figure 3**).

**Figure 3.**
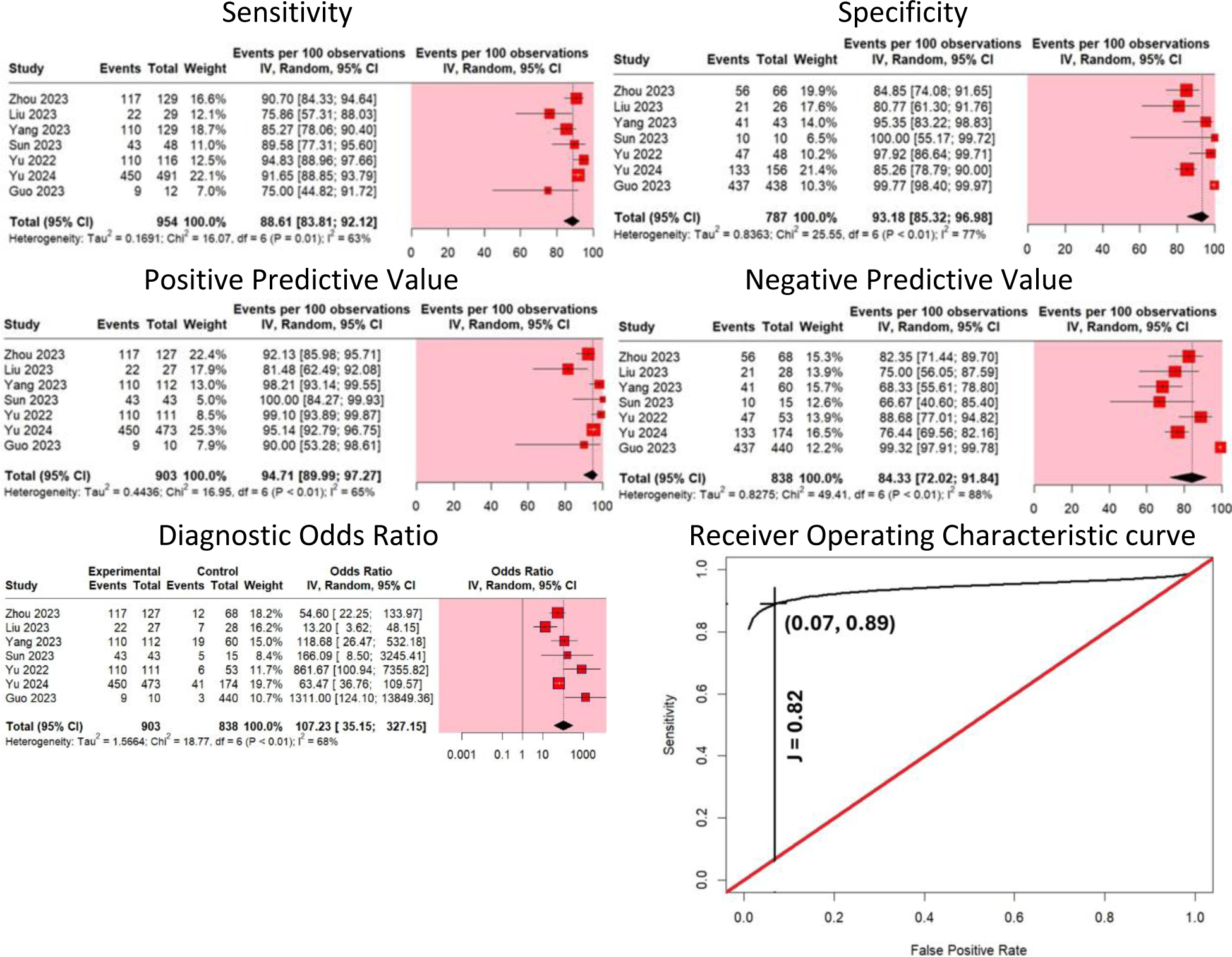
Diagnostic accuracy measures of nanopore sequencing in MTB detection.

*Publication bias*. Deek’s test was not performed since there were only 7 studies with calculated DORs at most (n<10).

### Detection of DR-MTB

Among 19 studies selected, 11 studies adopted a prospective cross-sectional design, while 7 used a retrospective cross-sectional approach (**Table 2**). MinION alone was predominantly used in these studies (14 out of 19), followed by GridION alone (3 out of 19), then the combination of both sequencing platforms (2 out of 19). These studies span publication years from 2017 to 2024. No study used Flongle or PromethION.

### Risk of bias and applicability assessment for DR-MTB

Among the 16 DR-MTB studies, two studies followed STARD recommendations^50^; one study stated the use of consecutive sampling^47^ and one study mentioned use of random sampling^41^. Further lack of clinical information about the samples contributed to the applicability concerns in drug resistance studies. Additionally, 6 out of the 16 DR-MTB studies employed a case-control design, amplifying the risk of bias related to the index test, reference test, and the sequencing of events in these investigations (**Figure 4**).

**Figure 4.**
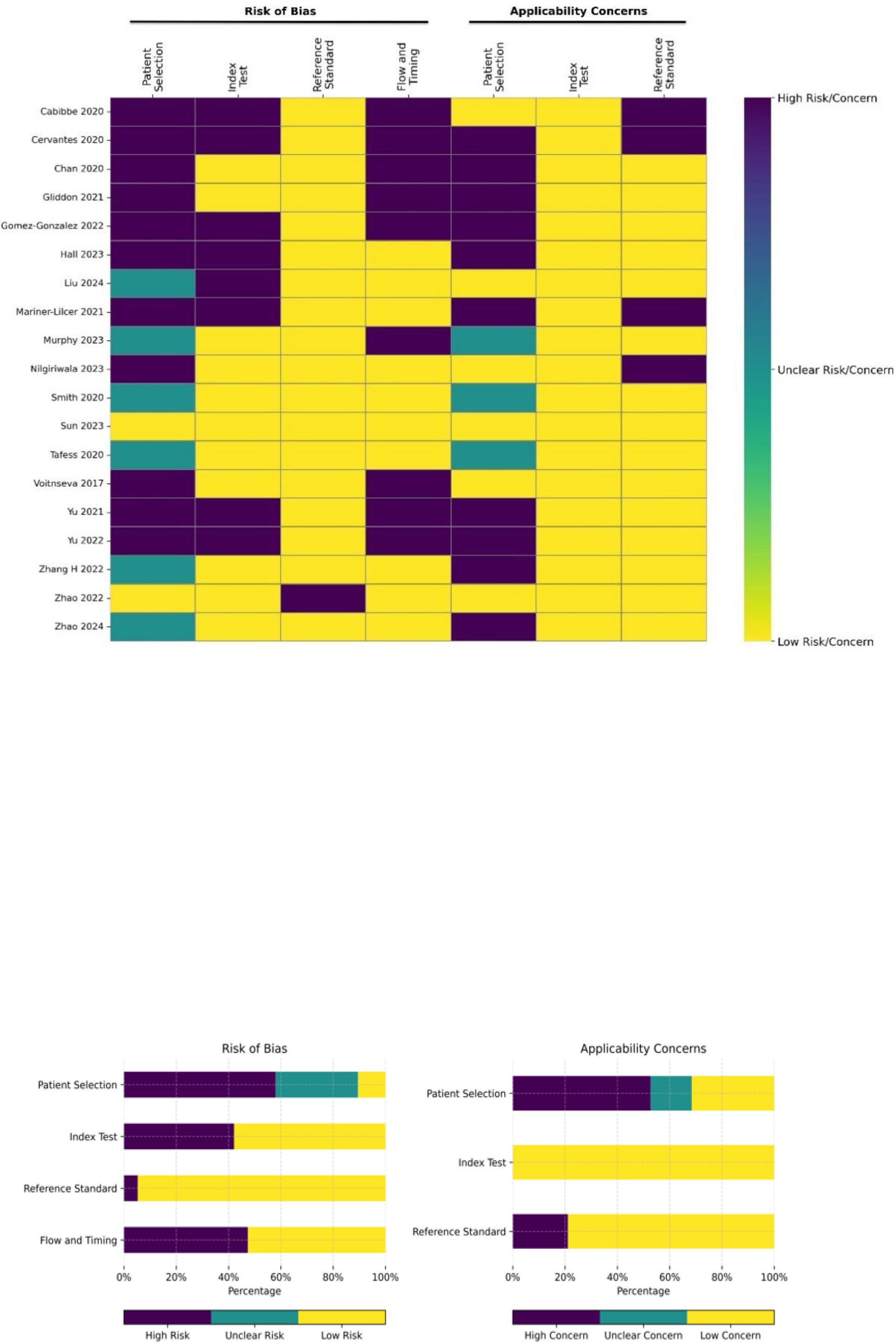
Heat maps showing risk of bias and applicability concerns among studies focusing on the detection of DR-MTB.

### Meta-analysis of studies on DR-MTB

*Heterogeneity test.* Our results showed that the logit-transformed sensitivity did not correlate with the logit-transformed false positive rate (1-specificity) across all drug resistance groups [Spearman coefficient range: -0.5 to 0.5 (*p-value range* = 0. 5 to 1)]. Higgin’s I2 of the diagnostic measures ranged from 0% to 87%.

*Evaluation of diagnostic accuracy.* The meta-analysis on detecting DR-MTB comprised 917 adult participants from five studies. Our results indicated that pooled sensitivity of nanopore sequencing was lower than its specificity in detecting any drug resistance; pooled sensitivity ranged from 77.99% to 93.66%, while the pooled specificity varied between 94.84% and 99.47% (**Table 3**). The pooled PPV ranged from 87.57% to 100% while the pooled NPV varied between 91.27% and 99.46% (**Table 3**).

**Table 3.**
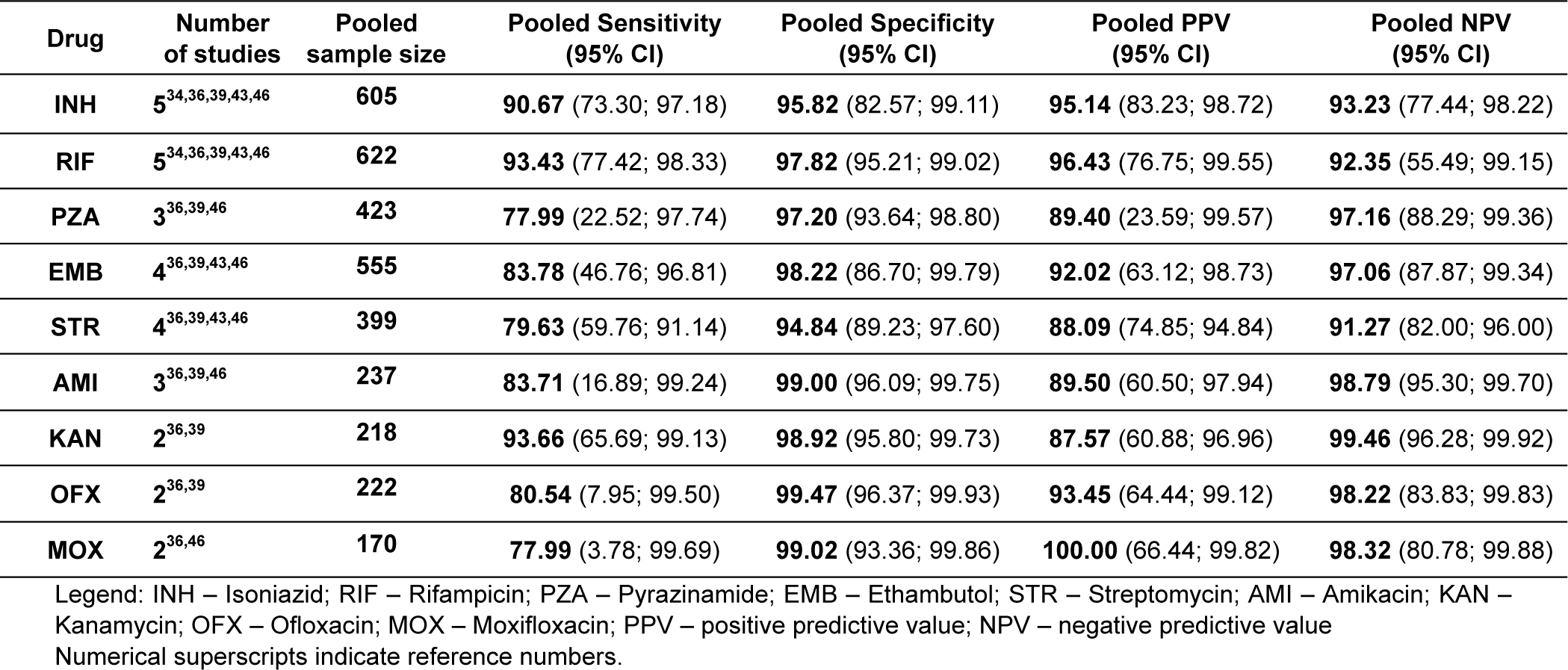
Summary of pooled diagnostic indices for nanopore sequencing in the detection of DR-MTB.

| Drug | Number of studies | Pooled sample size | Pooled Sensitivity (95% CI) | Pooled Specificity (95% CI) | Pooled PPV (95% CI) | Pooled NPV (95% CI) |
| --- | --- | --- | --- | --- | --- | --- |
| INH | 5<sup>34,36,39,43,46</sup> | 605 | 90.67 (73.30; 97.18) | 95.82 (82.57; 99.11) | 95.14 (83.23; 98.72) | 93.23 (77.44; 98.22) |
| RIF | 5<sup>34,36,39,43,46</sup> | 622 | 93.43 (77.42; 98.33) | 97.82 (95.21; 99.02) | 96.43 (76.75; 99.55) | 92.35 (55.49; 99.15) |
| PZA | 3<sup>36,39,46</sup> | 423 | 77.99 (22.52; 97.74) | 97.20 (93.64; 98.80) | 89.40 (23.59; 99.57) | 97.16 (88.29; 99.36) |
| EMB | 4<sup>36,39,43,46</sup> | 555 | 83.78 (46.76; 96.81) | 98.22 (86.70; 99.79) | 92.02 (63.12; 98.73) | 97.06 (87.87; 99.34) |
| STR | 4<sup>36,39,43,46</sup> | 399 | 79.63 (59.76; 91.14) | 94.84 (89.23; 97.60) | 88.09 (74.85; 94.84) | 91.27 (82.00; 96.00) |
| AMI | 3<sup>36,39,46</sup> | 237 | 83.71 (16.89; 99.24) | 99.00 (96.09; 99.75) | 89.50 (60.50; 97.94) | 98.79 (95.30; 99.70) |
| KAN | 2<sup>36,39</sup> | 218 | 93.66 (65.69; 99.13) | 98.92 (95.80; 99.73) | 87.57 (60.88; 96.96) | 99.46 (96.28; 99.92) |
| OFX | 2<sup>36,39</sup> | 222 | 80.54 (7.95; 99.50) | 99.47 (96.37; 99.93) | 93.45 (64.44; 99.12) | 98.22 (83.83; 99.83) |
| MOX | 2<sup>36,46</sup> | 170 | 77.99 (3.78; 99.69) | 99.02 (93.36; 99.86) | 100.00 (66.44; 99.82) | 98.32 (80.78; 99.88) |

Legend: INH – Isoniazid; RIF – Rifampicin; PZA – Pyrazinamide; EMB – Ethambutol; STR – Streptomycin; AMI – Amikacin; KAN – Kanamycin; OFX – Ofloxacin; MOX – Moxifloxacin; PPV – positive predictive value; NPV – negative predictive value Numerical superscripts indicate reference numbers.

Our results indicated that among first-line DST, accurate detection of pyrazinamide-resistance was most challenging in terms of sensitivity and PPV. Conversely, detection of rifampicin resistance was easiest for nanopore sequencing among first-line DST in terms of sensitivity and PPV. Pooled specificity of nanopore sequencing was highest in detecting ethambutol-resistance and lowest in isoniazid resistance. Pooled NPV, on the other hand, was highest in detecting pyrazinamide-resistance and lowest in rifampicin-resistance. Meanwhile, among second-line DST, the pooled sensitivity and NPV of nanopore sequencing were the highest but PPV was the lowest in detecting kanamycin-resistance. The highest specificity was noted in detecting ofloxacin resistance. Detection of streptomycin-resistance appeared to be more challenging; pooled specificity and NPV was at its lowest for MTB resistant to this aminoglycoside. Interestingly, pooled sensitivity was lowest, but pooled PPV was the highest for moxifloxacin-resistance. Resistance to ethionamide^39^, ciprofloxacin^36^, capreomycin^36^, and bedaquiline^43^ were each investigated by only one study; diagnostic accuracy measures are shown in **Table 4**. However, only one bedaquiline-resistant sample was detected in the sole study it was mentioned, making calculation of diagnostic accuracy measures impossible. Furthermore, no study mentioned about pretomanid or linezolid resistance. Among these three drugs, nanopore sequencing had the lowest diagnostic accuracy measures in detecting ethionamide resistance.

**Table 4.** Summary of diagnostic indices of nanopore sequencing in detecting DR-MTB resistant to drugs mentioned in only one study^36,39,43^.

| Drug | Sensitivity (95% CI) | Specificity (95% CI) | PPV (95% CI) | NPV (95% CI) | DOR (95% CI) | AUC |
| --- | --- | --- | --- | --- | --- | --- |
| ETH<sup>39</sup> | 50.00<br>(31.67; 68.33) | 97.30<br>(89.83; 99.32) | 86.67<br>(59.46; 96.64) | 84.71<br>(75.42; 90.91) | 36.00<br>(7.26; 178.59) | 0.99 |
| CIP<sup>36</sup> | 100.00<br>(66.44; 99.82) | 100.00<br>(94.40; 99.98) | 100.00<br>(66.44; 99.82) | 100.00<br>(94.40; 99.98) | 8943.00<br>(171.66; 465900.13) | 0.99 |
| CAP<sup>36</sup> | 100.00<br>(61.61; 99.78) | 100.00<br>(94.52; 99.98) | 100.00<br>(61.61; 99.78) | 100.00<br>(94.52; 99.98) | 7479.00<br>(142.64; 392156.18) | 0.99 |

Legend: ETH – ethionamide; CIP – ciprofloxacin; CAP – capreomycin; PPV – positive predictive value; NPV – negative predictive value; DOR – diagnostic odds ratio; AUC – area under the curve

The AUC indicated clinically acceptable performance across all DST studies, with values ranging from 0.85 to 0.990 (**Table 5**), observed for both first-line and second-line DST (**Figure 5**)^51^. AUC was excellent in most of DST investigated; it was lower than 0.9 only in detection of ethambutol and streptomycin resistance. The range of pooled DOR was remarkably wide; values varied from 106.94 to 8943.00 (**Table 5**). DOR was the lowest in detecting isoniazid and streptomycin resistance, and the highest in detecting pyrazinamide and moxifloxacin resistance.

**Figure 5.**
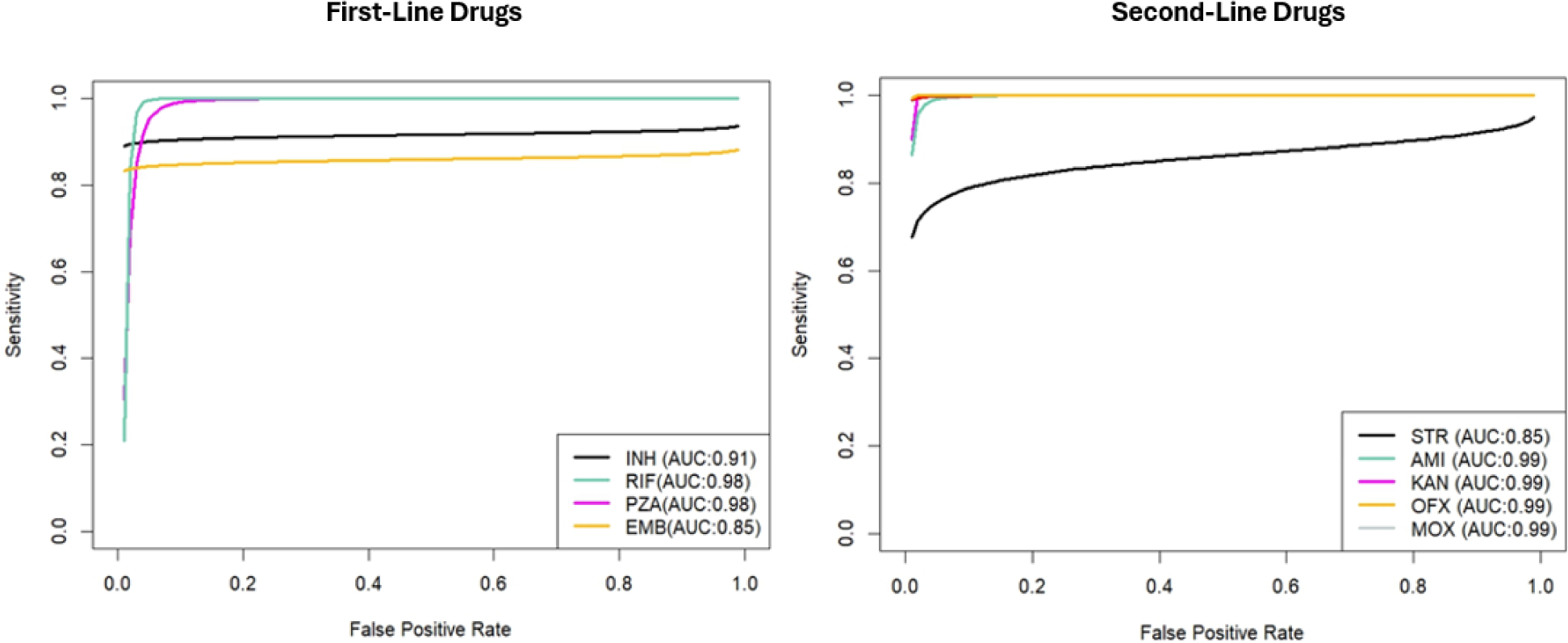
Summary ROC curve of nanopore sequencing in detection of DR-MTB resistant to first-line drugs. INH – Isoniazid; RIF – Rifampicin; PZA – Pyrazinamide; EMB – Ethambutol; and second-line drugs. STR – Streptomycin; AMI – Amikacin; KAN – Kanamycin; OFX – Ofloxacin; MOX – Moxifloxacin; CAP – Capreomycin.

**Table 5.** Diagnostic accuracy of nanopore sequencing in the detection of DR-MTB.

| Antibiotic | DOR<br>95% CI | AUC |
| --- | --- | --- |
| INH | **265.30** (17.80; 3953.16) | 0.91 |
| RIF | **386.97** (44.41; 3371.57) | 0.98 |
| PZA | **806.33** (5.67; 114649.22) | 0.98 |
| EMB | **744.07** (21.46; 25799.73) | 0.85 |
| STR | **106.94** (12.47; 917.27) | 0.85 |
| AMI | **625.53** (34.83; 11234.81) | 0.99 |
| KAN | **916.31** (88.25; 9514.06) | 0.99 |
| OFX | **815.34** (9.10; 73012.26) | 0.99 |
| MOX | **8943.00** (171.66; 465900.13) | 0.99 |

Legend: INH – Isoniazid; RIF – Rifampicin; PZA – Pyrazinamide; EMB – Ethambutol; STR – Streptomycin; AMI – Amikacin; KAN – Kanamycin; OFX – Ofloxacin; MOX – Moxifloxacin; DOR – diagnostic odds ratio; AUC – area under the curve

*Publication bias*. Deek’s test was not performed since there were only 4 studies with calculated DOR at most (n<10).

## DISCUSSION

Most of the studies aimed at evaluating only MTB detection by nanopore sequencing used GridION. In fact, GridION was used in all the studies included in this meta-analysis. Our study shows that GridION has high sensitivity [88.61%; 95% CI (83.81–92.12%)] and specificity [93.18%; 95% CI (85.32–96.98%)] in detecting MTB, indicating that this technology is accurate enough when compared to reference genotypic or phenotypic methods^52^. Its high PPV [94.71%; 95% CI (89.99–97.27%)] and moderately high NPV [84.31%; 95% CI (72.02–91.84%)] suggests its usefulness in clinical practice^52^ GridION also has an excellent discriminatory performance, as reflected by its large DOR [107.23; 95% CI (35.15–327.15)], which far exceeded the minimum standard (DOR = 10) for a very good test, its AUC (0.932) which exceeded 0.9, and its Youden index (J = 0.82), which exceeded 0.50^53-55^.

Most of the studies investigating DST by nanopore sequencing, including those considered in our meta-analysis, used minION. Estimates for pooled sensitivity were imprecise; a study using DNA extraction from sputum, instead of the usual DNA extracts from culture, recorded low sensitivities ranging from 0 to 33%^37^. Among first-line DST, nanopore sequencing performed best in detecting pyrazinamide and rifampicin resistance. Pyrazinamide had the highest DOR (806.33) and AUC (0.98) despite having the lowest sensitivity (77.99) and PPV (89.40). Rifampicin also had the highest AUC, tied with pyrazinamide, due to its sensitivity (93.43) and PPV (96.43) which were highest among first-line drugs. Isoniazid resistance has been a challenge, with the lowest DOR (265.30) and second to the lowest AUC (0.91), primarily due to its specificity (95.82), which was lowest. Similarly, ethambutol had the lowest AUC (0.85) despite having the highest specificity (98.22).

On the other hand, among second-line DST, nanopore sequencing excelled in detecting resistance to most second-line drugs, especially moxifloxacin. Moxifloxacin resistance had the highest DOR (8943.00), AUC (0.99) and PPV (100) despite having the lowest sensitivity (77.99). Notably, AUC was at its highest value (0.99) for all second-line drugs, except streptomycin. Streptomycin resistance detection had the lowest performance in DOR (106.94) and AUC (0.85) mainly due to its specificity (94.84) and NPV (91.27), which were also lowest. This may be attributed to still unknown resistance mechanisms, a finding consistent with previously reported conflicts with DST^39^. Furthermore, streptomycin resistance involves several genes (e.g., rpsL, rrs); the presence of multiple potential sites for mutations makes comprehensive detection more challenging. While streptomycin is a valuable option for the treatment of DR-TB, its usage is in decline due to the expanded acquisition and maintenance of resistant MTB isolates^56^.

Mathematically, DOR is directly related to AUC; the higher the DOR, the higher the AUC^53^. However, it has been shown that AUC is maximized when the study diagnostic odds ratios are homogeneous^57^. This explains why even though among first-line drugs, the DOR of ethambutol resistance detection is second to the highest [DOR 744.07; 95% CI (21.46; 25799.73)], it has the lowest AUC (0.85). Furthermore, the DOR of isoniazid is lowest [DOR 265.30; 95% CI (17.80; 3953.16)] but it has the second highest AUC (0.91). Indeed, the precision and not only the point estimate of DOR affects AUC. It is also notable that the heterogeneity of most diagnostic measures is remarkable (I2 > 50%); however, this is common with meta-analyses of diagnostic accuracy research^58^.

One of the primary weaknesses of nanopore sequencing technology, specifically MinION, in its early use, is its high error rate (20-35%)^59^. However, the change of assay chemistry from R7 to R9 by using an *Escherichia coli* pore protein CsgG, the shift from HMM-based base-caller to neural network-based base-caller, the refinement of statistical techniques to reduce structural noise, and the use of post-sequencing correction tools greatly improved the performance of nanopore sequencing technology, which is partly evidenced by the results of our study^60,61^. This study shows that nanopore sequencing technology is accurate and useful in the detection of both DS-MTB and DR-MTB strains in clinical specimens.

This study has several limitations, all stemming from the small number of studies included in the actual meta-analysis. The effects of variations in reference standard and age of participants, among other pertinent variables, on the diagnostic accuracy measures could be accounted for by subgroup analysis or meta-regression. These tests were not done since the number of studies is less than 10^62^. Publication bias was also not investigated due to the same reason^62^. Additional studies will also be needed to provide more reliable estimates of the diagnostic accuracy of nanopore sequencing. The lower 95% Cl estimates of DOR in detecting resistance to pyrazinamide (5.67), streptomycin (7.90), and ofloxacin (9.10) were found to be <10, which is the minimum DOR for a very good test. Even though this imprecision may cast uncertainty in the performance of nanopore sequencing in detecting MTB isolates resistant for these three drugs, the use of nanopore sequencing for MTB DST is only emerging. More studies will also provide more reliable estimates of the diagnostic accuracy of nanopore sequencing in detecting resistance to ethionamide, ciprofloxacin, and capreomycin. In this context, it is notable that nanopore sequencing, just like other sequencing platforms, fares relatively poorly in detecting ethionamide resistance, which could be explained by the presence of unknown resistance mechanisms for ethionamide, which is also one of the primary reasons for overall conflicts with DST^39^. Furthermore, diagnostic accuracy studies on bedaquiline, pretomanid, and linezolid should be of top priority since statistical studies about these drugs are absent.

## CONCLUSION

Nanopore sequencing GridION has high sensitivity and specificity, high PPV, moderately high NPV, and excellent DOR and AUC in detecting MTB. MinION excels in detecting pyrazinamide and rifampicin resistance and underperforms in detecting isoniazid and ethambutol resistance when compared to current phenotypic DST methods. However, additional studies are required to provide more precise estimates for MinION’s sensitivity in detecting drug-resistance and DOR in detecting resistance to pyrazinamide, streptomycin, and ofloxacin. Furthermore, studies on detecting resistance to bedaquiline, pretomanid, and linezolid are lacking.

## Author roles

All authors meet the journal’s criteria for authorship. Individual contributions/author roles are listed below.

## Authors Contribution/s

Timothy Hudson David Culasino Carandang: conceptualization, visualization, methodology, statistical analysis, validation, data curation, writing-original draft, writing-review and editing

Dianne Jaula Cunanan: visualization, methodology, validation, data curation, writing-original draft, writing-review and editing

Gail S. Co: methodology, validation, data curation, writing-original draft, writing-review and editing

John David Pilapil: visualization, methodology, validation, data curation, writing-review and editing

Juan Ignacio Garcia: writing-review and editing

Blanca I. Restrepo: writing-review and editing

Marcel Yotebieng: writing-review and editing

Jordi B. Torrelles: writing-review and editing

Kin Israel Notarte: conceptualization, visualization, methodology, validation, data curation, writing-review and editing

## Data Availability

All data produced in the present work are contained in the manuscript

## SUPPLEMENTARY MATERIALS

**Supplementary Table 1.**
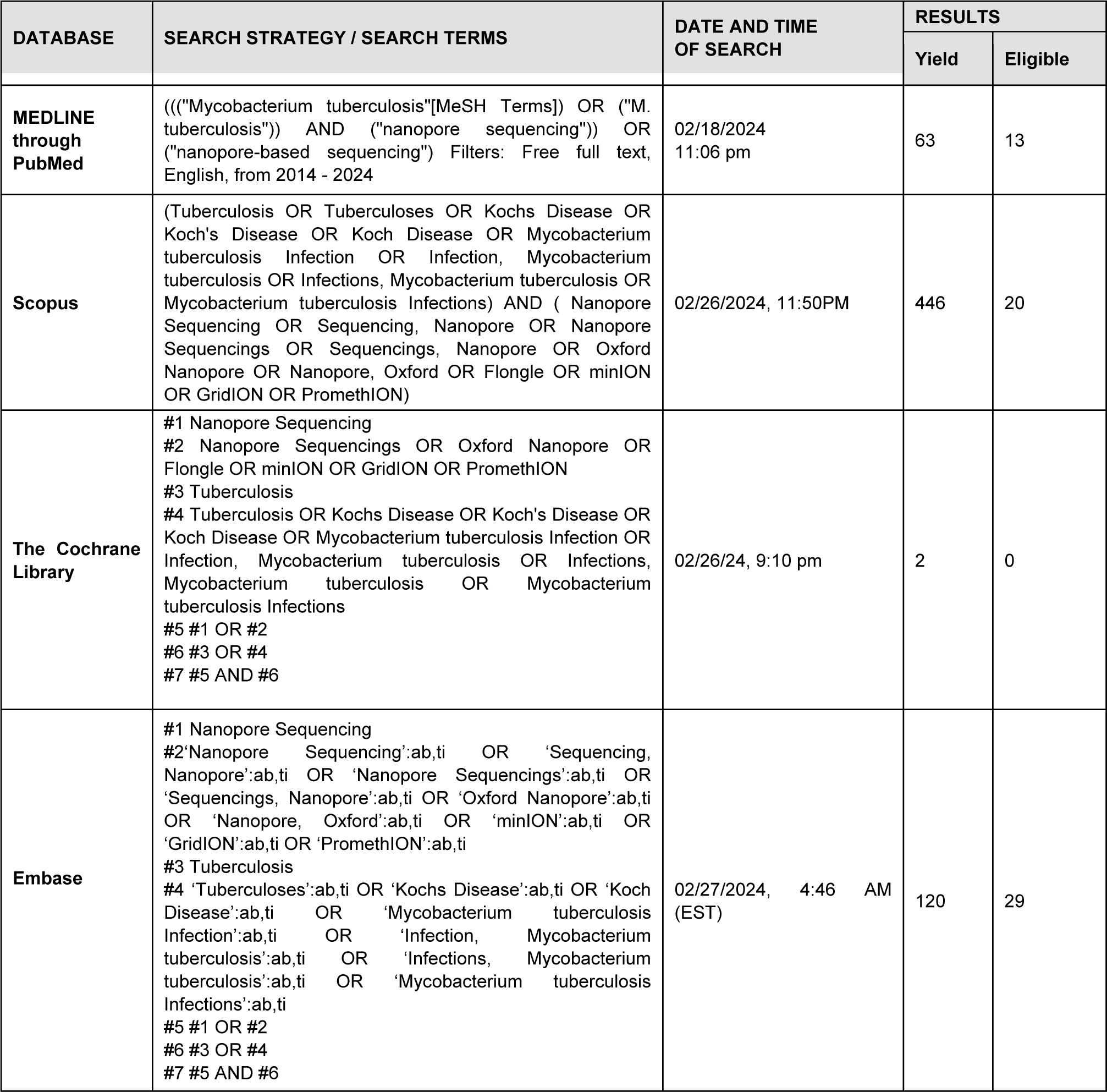
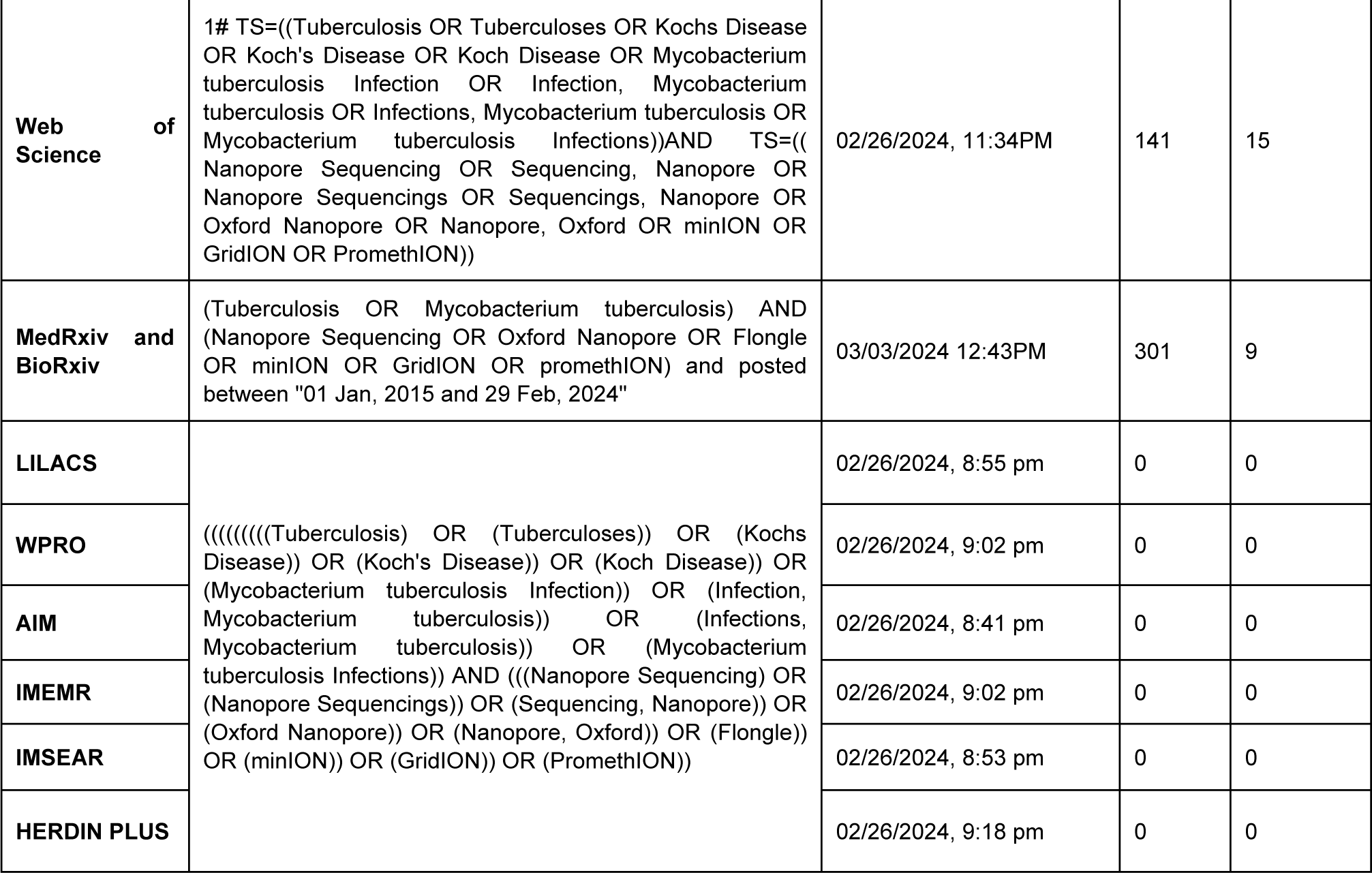
Full Search Strategy for MTB Detection Studies.

**Supplementary Table 2.** Full Search Strategy for MTB Drug Susceptibility Studies.

| DATABASE | SEARCH STRATEGY / SEARCH TERMS | DATE AND TIME OF SEARCH | RESULTS |  |
| --- | --- | --- | --- | --- |
|  |  |  | Yield | Eligible |
| MEDLINE through PubMed | ((isoniazid OR rifampicin OR pyrazinamide OR ethambutol OR streptomycin OR amikacin OR ofloxacin OR moxifloxacin OR capreomycin OR ciprofloxacin OR ethionamide OR kanamycin OR bedaquiline OR pretomanid OR linezolid) AND (resistant OR resistance)) AND (Tuberculosis OR Tuberculoses OR Kochs Disease OR Koch's Disease OR Koch Disease OR Mycobacterium tuberculosis Infection OR Infection, Mycobacterium tuberculosis OR Infections, Mycobacterium tuberculosis OR Mycobacterium tuberculosis Infections)) AND ( Nanopore Sequencing OR Sequencing, Nanopore OR Nanopore Sequencings OR Sequencings, Nanopore OR Oxford Nanopore OR Nanopore, Oxford OR Flongle OR minION OR GridION OR PromethION) | 05/09/2024<br>8:45 pm | 13 | 7 |
| Cochrane Library |  | 05/09/2024<br>11:14 pm | 0 | 0 |
| Embase | #1 Nanopore Sequencing<br>#2'Nanopore Sequencing':ab,ti OR 'Sequencing, Nanopore':ab,ti OR 'Nanopore Sequencings':ab,ti OR 'Sequencings, Nanopore':ab,ti OR 'Oxford Nanopore':ab,ti OR 'Nanopore, Oxford':ab,ti OR 'minION':ab,ti OR 'GridION':ab,ti OR 'PromethION':ab,ti<br>#3 Tuberculosis<br>#4 'Tuberculoses':ab,ti OR 'Kochs Disease':ab,ti OR 'Koch Disease':ab,ti OR 'Mycobacterium tuberculosis Infection':ab,ti OR 'Infection, Mycobacterium tuberculosis':ab,ti OR 'Infections, Mycobacterium tuberculosis':ab,ti OR 'Mycobacterium tuberculosis Infections':ab,ti<br>#5 drug resistant<br>#6 'isoniazid':ab,ti OR 'rifampicin':ab,ti OR 'pyrazinamide':ab,ti OR 'ethambutol':ab,ti OR 'streptomycin':ab,ti OR 'amikacin':ab,ti OR 'ofloxacin':ab,ti OR 'moxifloxacin':ab,ti OR 'capreomycin':ab,ti OR 'ciprofloxacin':ab,ti OR 'ethionamide':ab,ti OR 'kanamycin':ab,ti OR 'bedaquiline':ab,ti OR 'pretomanid':ab,ti OR 'linezolid':ab,ti<br>#7 'resistant':ab,ti OR 'resistance':ab,ti<br>#8 #1 OR #2<br>#9 #3 OR #4<br>#10 #6 AND #7<br>#11 #5 OR #10<br>#12 #8 AND #9<br>#13 #11 AND #12 | 05/10/2024<br>5:16 pm | 25 | 11 |
| Web of Science | 1# TS=((Tuberculosis OR Tuberculoses OR Kochs Disease OR Koch's Disease OR Koch Disease OR Mycobacterium tuberculosis Infection OR Infection, Mycobacterium tuberculosis OR Infections, Mycobacterium tuberculosis OR Mycobacterium tuberculosis Infections))AND TS=((isoniazid OR rifampicin OR pyrazinamide OR ethambutol OR streptomycin OR amikacin OR ofloxacin OR moxifloxacin OR capreomycin OR ciprofloxacin OR ethionamide OR kanamycin OR bedaquiline OR pretomanid OR linezolid))AND TS=(( Nanopore Sequencing OR Sequencing, Nanopore OR Nanopore Sequencings OR Sequencings, Nanopore OR Oxford Nanopore OR Nanopore, Oxford OR minION OR GridION OR PromethION)) | 05/10/2024 12:04pm | 13 | 8 |
| Scopus | ((isoniazid OR rifampicin OR pyrazinamide OR ethambutol OR streptomycin OR amikacin OR ofloxacin OR moxifloxacin OR capreomycin OR ciprofloxacin OR ethionamide OR kanamycin OR bedaquiline OR pretomanid OR linezolid) AND (resistant OR resistance)) AND (Tuberculosis OR Tuberculoses OR Kochs Disease OR Koch's Disease OR Koch Disease OR Mycobacterium tuberculosis Infection OR Infection, Mycobacterium tuberculosis OR Infections, Mycobacterium tuberculosis OR Mycobacterium tuberculosis Infections) AND (Nanopore Sequencing OR Sequencing, Nanopore OR Nanopore Sequencings OR Sequencings, Nanopore OR Oxford Nanopore OR Nanopore, Oxford OR minION OR GridION OR PromethION) | 10 May 2024 12:10pm | 161 | 14 |
| medRxiv and bioRxiv | ((bedaquiline) AND (resistance)) AND (Tuberculosis) AND (Nanopore Sequencing OR Flongle OR minION OR GridION OR PromethION) and posted between "01 Jan, 2014 and 29 Feb, 2024" | 05/09/2024 11:07 PM | 14 | 3 |
| LILACS | ((isoniazid OR rifampicin OR pyrazinamide OR ethambutol OR streptomycin OR amikacin OR ofloxacin OR moxifloxacin OR capreomycin OR ciprofloxacin OR ethionamide OR kanamycin OR bedaquiline OR pretomanid OR linezolid) AND (resistant OR resistance)) AND (Tuberculosis OR Tuberculoses OR Kochs Disease OR Koch's Disease OR Koch Disease OR Mycobacterium tuberculosis Infection OR Infection, Mycobacterium tuberculosis OR Infections, Mycobacterium tuberculosis OR Mycobacterium tuberculosis Infections)) AND ( Nanopore Sequencing OR Sequencing, Nanopore OR Nanopore Sequencings OR Sequencings, Nanopore OR Oxford Nanopore OR Nanopore, Oxford OR Flongle OR minION OR GridION OR PromethION) | 05/09/2024, 10:30 pm | 0 | 0 |
| WPRO |  | 02/26/2024, 9:02 pm | 0 | 0 |
| AIM |  | 02/26/2024, 8:41 pm | 0 | 0 |
| IMEMR |  | 02/26/2024, 9:02 pm | 0 | 0 |
| IMSEAR |  | 02/26/2024, 8:53 pm | 0 | 0 |
| HERDIN PLUS |  | 02/26/2024, 9:18 pm | 0 | 0 |

